# A mixed method study to assess behavioral and social predictors of parent/caregiver’s intention to vaccinate their children against COVID-19 disease in an Indian state marked by significant health disparities

**DOI:** 10.1101/2023.10.05.23296592

**Authors:** Tulika Singh, Sanjay Kumar, Setu Sinha, Varsha Singh

## Abstract

**Background:** Parents/caregivers are the key decision-makers for child’s health care including vaccination. Vaccine hesitancy along with lagging full immunization coverage for childhood vaccination in India, affect child health outcome and will affect covid-19 vaccine uptake in children. It is important to understand behavioral and social factors surrounding childhood COVID-19 vaccination to design appropriate interventions to improve uptake.

**Methods:** A mixed-method approach combining quantitative and qualitative method was undertaken. A cross sectional survey of parents/caregivers of children aged less than 18 years residing in the state was carried out to find the prevalence and predictors of parent/ caregiver’s intention to vaccinate against COVID-19 disease. Semi-structured interviews were carried out to find facilitating and barrier factors for childhood COVID-19 vaccination.

**Result:** Out of 9904 study participants, 73.4% had intention to vaccinate. Parent/caregiver’s education and occupation, marital status, family type, family income, co-morbidity and previous COVID-19 infection in family, childhood vaccination under NIS, were found to be significantly associated. The likelihood of intention to vaccinate children against COVID-19 disease was greater among parents/caregivers aged 18-29 years (OR=2.631, 95% CI [1.733- 3.995], illiterate parents/caregivers (OR=3.037, 95% CI [2.319-3.977], prior COVID-19 infection in family (OR=1.595, 95% CI [1.432-1.821], and children’s prior vaccinations under NIS (OR=1.251, 95% CI [1.218-1.289]. In qualitative part, forty-five semi-structured interviews were conducted. The majority of intending parents gave vaccine effectiveness, increased immunity, high infection risk, herd immunity, and medical recommendations as reasons. Parents who refused mentioned inadequate data, adverse effects, beliefs, safety, and inconvenience as reasons. Effectiveness, and safety, long-term effects, and the short testing period were among the concerns of hesitant parents.

**Conclusion:** In order to promote COVID-19 vaccination among children, we need to address barriers, facilitators and behavioral determinants of parents/caregivers identified in this study and have targeted strategies for them.

## Introduction

Since March 2020, COVID-19 pandemic caused by severe acute respiratory syndrome coronavirus 2 (SARS-CoV-2), has had a profound impact on the world. It has impacted almost every aspect of daily life. The pandemic has resulted in far-reaching social, economic, and health consequences, such as loss of lives, increased hospitalizations, prompted mental health crisis and job losses.^1,2,3^ There have been efforts to create and distribute effective vaccines to stop the spread of COVID-19 since the pandemic began. An exceptional accomplishment has been achieved through the mobilization of global resources, resulting in the development of safe and effective vaccines within an unprecedentedly short time frame.^4,5^

Vaccines are one of the most effective ways to prevent the spread of infectious diseases and have played a critical role in reducing the incidence of many infectious diseases globally. In modern healthcare, vaccines hold immense public health value as they are a cost-effective means of promoting global welfare.^6–8^ They have been recognized as one of the ten most significant public health achievements of the 20th and 21st centuries.^9^ The approved COVID- 19 vaccines have been scientifically proven to be highly effective and safe in preventing moderate to severe forms of the disease.^10–12^

However, the development of a vaccine is not sufficient, as modelling suggests that up to 80% of the population needs to receive a vaccine that is 70% effective in order to end the pandemic without additional non-pharmaceutical interventions.^13^ Vaccine uptake relies not only on adequate production and distribution, but also on high levels of vaccine acceptance among the general public.^14^ The WHO has listed vaccine hesitancy, defined as the delay in acceptance or refusal of vaccines despite availability of vaccination services, as one of the top ten threats to global health, even prior to the current COVID-19 pandemic.^15,16^ Population with more vaccine hesitancy has more chance of contracting and spreading the disease.

Despite the global spread, epidemiology and clinical presentation of COVID-19 in children remains largely unclear. The perceived severity of COVID-19 in children is less than that in the adults, but long-term serious complications of COVID-19 in children have been increasingly reported including long-COVID symptoms and multisystem inflammatory syndrome in children (MIS-C). These complications could be prevented by COVID-19 vaccines. As a result, every child should be managed to receive the vaccine timely.^17^

India launched vaccination against COVID-19 on January 16, 2021 and till date around 85% of eligible population is fully vaccinated with two doses of Covid-19 vaccines, and only 21% of the population has received booster dose.^18^ Bihar is lagging behind in vaccine uptake as analysis of persons aged 18-44 years and 45 years and above showed percentage fully vaccinated at ∼54% and ∼55% respectively. Keeping in view of the global surge of COVID-19 cases, detection of Omicron variant, scientific evidence, global practices and the inputs/suggestions of ‘COVID-19 Working Group of National Technical Advisory Group on Immunization (NTAGI)’ as well as of ‘Standing Technical Scientific Committee (STSC)’ of NTAGI, the Government of India decided to further refine the scientific prioritization & coverage of COVID-19 vaccination and introduced COVID-19 Vaccination of children in the age-group of 15-18 years from 3rd January 2022.^19^ However, the rate of vaccine acceptance among children is still lower with only 38% of population in the age group of 12-17 years fully vaccinated. Vaccine hesitancy along with India’s lagging full immunization coverage for children affect child health outcome and will affect covid-19 vaccine uptake in children Parents/caregivers are the key decision-makers for child’s health care including vaccination. It is important to consider how personal beliefs, customs, values, and lifestyle affect a person’s decision surrounding their as well as children’s healthcare. Therefore, understanding what factors impact their decision-making prior to COVID-19 vaccine roll-out is critical for adequate uptake in children. A limited number of cross-sectional studies have been carried outside India^20–23^, and as per our knowledge there is non-availability of similar studies in India which have studied predictors for the same. Bihar has been lagging behind in vaccination specially in the rural areas where vaccine hesitancy is quite high and associated with rumors and myths^24^.

We planned a sequential explanatory mixed-method study, with cross-sectional questionnaire-based survey to determine the prevalence of intention to vaccinate among parents/caregivers for pediatric COVID-19 vaccination, followed by semi-structured interviews to explore the facilitating and barrier factors, and to find solutions to improve pediatric COVID-19 vaccine uptake in the state of Bihar.

## Materials and Methods

**Study population**: Parents/Caregivers of children aged less than 18 years residing in Bihar

**Study period**: 6 months (April 2022- September 2022)

**Study design**: Mixed-method approach combining quantitative (Cross-sectional) and qualitative method (Semi-structured interviews).

**Inclusion criteria**: Adult parents/caregivers of children under 18 years of age and resident of Bihar

**Exclusion criteria:** Individuals with serious physical/mental health problems or physically disabled

### Phase-1 Quantitative (Cross-sectional study)

#### Study setting & Period

A community-based mixed-method study was carried out among parents of children aged less than 18 years residing in the state of Bihar. The state is divided into 38 districts and an online questionnaire was utilized in an attempt to collect data from every district during the ongoing Covid-19 pandemic. The study was conducted during the month of April to August 2022.

#### Sample Size and Sampling Technique

The minimum sample size was calculated to be **9604**, based on the conservative assumption that the acceptability rate (p) in parents is 50% with a 1% margin of error (M) and a confidence interval of 95%, using the formula n= Z_α/2_^2^ *p*(1-p) / M^2^ Due to limitations in doing face-to-face survey during the active COVID-19 pandemic, online cross-sectional questionnaire-based survey utilizing the social media platform and random digit dialing to survey respondents was carried out. The study details were advertised outlining the inclusion and exclusion criteria for participation and included hyperlink to the survey for participants. The paid promotion feature of social media platforms to target the survey at eligible participants was also utilized. Repeated reminders were sent to the non-responders. The principals of schools in the state were reached out via email, to disseminate the information to parents/caregivers, to obtain recruitment. Efforts were also made to include parents/caregivers who lacked internet access to ensure broader participation by providing them with hardcopies of the questionnaire, and the enumerators ensured to comply with health protocols in the midst of the pandemic. Internal validity was maintained by enforcing good data management practices.

#### Data Collection & Study Procedure

Respondents were provided with study details before starting the survey, explaining all aspects of participation. Participation in the study was voluntary. Anonymity and confidentiality of respondents was maintained and participants provided informed consent to be included in the study.

A bilingual, self-administered anonymous questionnaire was designed and pretested to ascertain comprehensibility and time taken before the final draft was made available to the participants. The online version of the final questionnaire took roughly 10 minutes to complete. Information on behavioral and social variables like age, gender, education, occupation, household income, history of COVID-19 disease in the family, vaccination status of parents/caregivers, and number and age of children were included. Parents/caregivers were also asked whether their children had received vaccinations under National Immunization Schedule (NIS) and intention to vaccinate their children against SARS-CoV-2. After making necessary modification to SAGE Vaccine hesitancy survey tool for assessing the intention to vaccinate, it was administered to participants.

### Phase 2: Qualitative (Semi-structured in-depth interview)

On completing the survey, participants were asked if they were willing to take part in a follow-up in-depth interview. Parents who answered yes, no as well as those who answered not sure for the question “If a COVID-19 vaccine is available to your child, will you get it?” were purposively selected. Recruitment stopped when data saturation was achieved (i.e., the point at which no new data was expressed). In total,45 interviews were conducted.

The aim was to identify facilitating factors among parents/caregivers who accepted vaccination and to explore barrier factors among those who refused/ were hesitant towards COVID-19 vaccination. To assist the interviewers in conducting semi-structured interviews with the participants, the research team developed in-depth interview guides based on pertinent vaccine hesitancy literature. These guides consisted of open-ended questions. During semi-structured interviews, researchers asked the same questions systematically to all participants while retaining the option to probe further by asking additional questions related to emerging themes during the interview. By using qualitative research methods, we were able to gain a more profound understanding of the decision-making process of parents/caregivers regarding COVID-19 vaccination for their children The interviewers possessed postgraduate degrees, were trained in qualitative research, and had fluency in the local language. We conducted the interviews over the phone in the local language, and each interview lasted between 30 to 40 minutes. Before beginning the interview, we obtained approval and spent the first 5 minutes explaining the purpose and motive of the study. We then proceeded to explore the participants’ perceptions of vaccination for themselves and their children. All information provided by the informant was kept strictly confidential. All the interviews were audio-recorded with consent. At the end of the interview, a summary was presented to the parents/caregivers for validation of the data collected. Transcription was carried out using a verbatim format within 2 days of data collection to prevent information loss.

### Data Analysis

Data were entered into Microsoft Excel and analysis was done using SPSS version 16, International Business Machines Corp., New York. The continuous variables such as age were summarized as mean (SD) and categorical as percentage/proportion. The prevalence of vaccine hesitancy was summarized as proportion with 95% confidence interval. Chi-square test/Fisher exact test was used to find the association between behavioral & socio-demographic factors and COVID-19 vaccine intention. Regression analyses were performed to assess the predictors of vaccine intention. A p<0.05 was considered to be statistically significant.

The qualitative data were analyzed with thematic analysis. Thematic analysis included, familiarization, coding, generating, reviewing and refining themes and attaching illustrative quotes to it. Initially, the entire dataset was transcribed verbatim and read twice to gain familiarity with the data. The unit of analysis was the participants’ statements. The next step involved coding the transcripts, which was done at the beginning of the study using a deductive method. As the research progressed, an inductive approach was also used during the review process. Descriptive manual content analysis was performed to derive the categories and codes from the collected data. The principal investigator validated the data and two other investigators reviewed it to minimize subjective interpretation. Constant comparative analysis was performed to ensure the credibility and reliability of the data. After refining and naming themes, illustrative quotes were attached. The Consolidated criteria for Reporting Qualitative Research checklist was used as guidelines for reporting items of interest in the presentation of qualitative results. This checklist ensured that the research findings were presented in a clear and transparent manner.

### Ethical considerations

The protocol of this study was approved by the Institutional Ethics Committee of IGIMS, Patna vide letter no.212/IEC/IGIMS/2021. All participants were informed about the objectives and purpose of this study and were asked to provide their voluntary consent for participation. Informed consent was requested on the introductory online questionnaire page before survey enrolment and only by selecting the consent option were respondents allowed access to survey questions. No incentives were offered to participants for enrolment. All personal information was kept confidential and the interviews were audio-recorded with consent.

## Results

### Quantitative part

In total 10809 parents/caregivers of children aged less than 18 years filled out the questionnaire for this study. Out of which 9904 were included in this study after checking for completeness of questionnaire and data cleaning (91.4% response rate).

The behavioral and sociodemographic features of the participants and its association with parents/caregivers’ intention to vaccinate their children against COVID-19 disease are included in Table 1. The mean age was found to be 37.09 (±8.83) and two third participants were male (65.24%). In educational attainment, one-third of the participants were graduate (35.15%) and less than 5% were illiterate. Majority of the participants worked in private sector (42.76%) and less than one fifth were either self-employed or working in government sector. More than half of the participants belonged to joint family (59.67%). More than three fourth of study participants did not report any co-morbidity (83.3%) and around one fifth had been infected with Covid-19 disease (21.5%).

**Table 1:** Association between parent’s intention to vaccinate their children against COVID-19 vaccination and behavioral and social factors.

| Variables | COVID-19 Vaccination for children |  | p value |
| --- | --- | --- | --- |
|  | Yes | No |  |
| **Age** |  |  |  |
| 18-29 | 1351 (66.7%) | 675 (33.3%) | 0.000 |
| 30-44 | 4464 (74.2%) | 1550 (25.8%) |  |
| 45-59 | 1318 (78.2%) | 366 (21.8%) |  |
| ≥60 | 133 (73.9%) | 47 (26.1) |  |
| **Gender** |  |  |  |
| Male | 4648 (71.9%) | 1813 (28.1%) | 0.000 |
| Female | 2606 (76.0%) | 823 (24.0%) |  |
| Prefer not to Say | 12 (85.7%) | 2 (14.3%) |  |
| **Education** |  |  |  |
| Illiterate | 148 (48.1%) | 160 (51.9%) | 0.000 |
| Primary | 499 (61.1%) | 318 (38.9%) |  |
| Secondary | 1229 (76.4%) | 380 (23.6%) |  |
| Senior secondary | 1257 (70.6%) | 523 (29.4%) |  |
| Graduate | 2578 (74.1%) | 903 (25.9%) |  |
| Post Graduate | 1014 (82.0%) | 222 (18.0%) |  |
| Professional | 541 (80.4%) | 132 (19.6%) |  |
| **Occupation** |  |  |  |
| Government | 1080 (73.6%) | 387 (26.4%) | 0.001 |
| Private | 3171 (74.9%) | 1064 (25.1%) |  |
| Self-employed | 1410 (73.7%) | 503 (26.3%) |  |
| Home maker | 1193 (69.8%) | 516 (30.2%) |  |
| Unemployed | 412 (71.0%) | 168 (29.0%) |  |
| **Marital status** |  |  |  |
| Married | 6701 (73.2%) | 2459 (26.8%) | 0.000 |
| Unmarried | 381 (87.6%) | 54 (12.4%) |  |
| Widow/ Widower | 139 (61.0%) | 89 (39.0%) |  |
| Divorced/ Separated | 45 (55.6%) | 36 (44.4%) |  |
| Religion |  |  |  |
| Hinduism | 6108 (77.8%) | 1741 (22.2%) | 0.000 |
| Islam | 707 (58.5%) | 502 (41.5%) |  |
| Christianity | 196 (51.3%) | 186 (48.7%) |  |
| Sikhism | 134 (51.9%) | 124 (48.1%) |  |
| Others | 121 (58.7%) | 85 (41.3%) |  |
| Family type |  |  |  |
| Joint | 4031 (68.2%) | 1879 (31.8%) | 0.000 |
| Nuclear | 3235 (81.0%) | 759 (19.0%) |  |
| Family income |  |  |  |
| ≤18604 | 3505 (75.1%) | 1162 (24.9%) | 0.000 |
| 18605-49730 | 2662 (66.3%) | 1351 (33.7%) |  |
| ≥ 49731 | 1099 (89.8%) | 125 (10.2%) |  |
| Co-morbidity |  |  |  |
| Yes | 592 (76.0%) | 187 (24.0%) | 0.014 |
| No | 6062 (73.5%) | 2186 (26.5%) |  |
| Not Aware | 612 (69.8%) | 265 (30.2%) |  |
| Infected covid |  |  |  |
| Yes | 1602 (75.2%) | 529 (24.8%) | 0.033 |
| No | 5664 (72.9%) | 2109 (27.1%) |  |
| Received covid vaccination |  |  |  |
| Yes | 7149 (73.7%) | 2547 (26.3%) | 0.000 |
| No | 117 (56.3%) | 91 (43.7%) |  |
| Children received childhood vaccines under NIS |  |  |  |
| Yes | 6781 (76.8%) | 2044 (23.2%) | 0.000 |
| No | 269 (52.3%) | 245 (47.7%) |  |
| Not sure | 216 (38.2) | 349 (61.8%) |  |

\*p<0.05, NIS=National Immunization Schedule

In our study 8230 (83.1%) parents/caregivers reported that their children have received childhood vaccination under NIS. More than half of the parents/caregivers were a little concerned about their children getting COVID-19 disease.

Out of 9904 study participants, 73.4% had intention to vaccinate their children against covid- 19 disease in the state of Bihar, 15.0% answered no and 11.6% were not sure about getting their children vaccinated against COVID-19. Education, occupation of respondent, marital status, family type, family income, co-morbidity, previous Covid infection, previous childhood vaccination status was found to be significantly associated with intention to vaccinate children against Covid-19 disease. [Table 1]

Results of binary logistics regression analysis of parent/ caregiver’s intention to vaccinate their children against COVID-19 disease and their predictors are presented in Table 2. Respondents aged 18-29 years demonstrated significantly higher odds (OR = 2.631, 95% CI [1.733-3.995], p < 0.001) of intending to vaccinate their children against COVID-19 compared to those aged ≥60 years. Illiterate parents/caregivers had 3.04 times higher odds (OR = 3.037, 95% CI [2.319- 3.977], p < 0.001) of intending to vaccinate compared to those with graduate & above education. Among different occupation categories, private sector employees (OR=0.769, 95% CI [0.609-0.971], p=0.027) and self-employed (OR=0.746, 95% CI [0.584-0.954], p=0.020) were less likely to express intent to vaccinate. Those belonging to joint family had 1.81 times higher odds (OR=1.814, 95% CI [1.630-2.020], p<0.001).

**Table 2:** Binary Logistics Regression Analysis of Parent/ caregivers intention to vaccinate their children against covid-19 disease and their predictors.

| Variable | Intended to vaccinate children |  |
| --- | --- | --- |
|  | Odds Ratio (95% CI) | p-value |
| Parent/caregiver's Age (in years) |  |  |
| 18-29 | 2.631 (1.733-3.995) | .000 |
| 30-44 | 1.828 (1.215-2.750) | .004 |
| 45-59 | 1.432 (0.945-2.170) | .090 |
| ≥60 | Reference |  |
| Parent/caregiver's Gender |  |  |
| Male | 3.525 (0.625-19.880) | .153 |
| Female | 2.164 (0.384-12.204) | .382 |
| Prefer not to Say | Reference |  |
| Parent/caregiver's Education |  |  |
| Illiterate | 3.037 (2.319-3.977) | .000 |
| Primary | 1.925 (1.616-2.294) | .000 |
| Secondary | 1.063 (0.914-1.236) | .430 |
| Senior secondary | 1.174 (1.026-1.343) | .019 |
| Graduate & above | Reference |  |
| Parent/caregiver's occupation |  |  |
| Government | .967 (0.746-1.254) | .800 |
| Private | .769 (0.609-0.971) | .027 |
| Self-employed | .746 (0.584-0.954) | .020 |
| Home maker | 1.239 (0.954-1.609) | .109 |
| Unemployed | Reference |  |
| Parent/caregiver's Marital status |  |  |
| Married | .516 (0.310-0.858) | .011 |
| Unmarried | .123 (0.067-0.226) | .000 |
| Widow/ Widower | .870 (0.480-1.575) | .645 |
| Divorced/ Separated | Reference |  |
| Religion |  |  |
| Hinduism | 1.691 (1.504-1.949) | .022 |
| Islam | 1.498 (1.072-2.094) | .018 |
| Christianity | 1.907 (1.306-2.782) | .001 |
| Sikhism | 1.543 (1.029-2.314) | .036 |
| Others | Reference |  |
| Type of Family |  |  |
| Joint | 1.814 (1.630-2.020) | .000 |
| Nuclear | Reference |  |
| Family Income (INR) |  |  |
| ≤18604 | 1.853 (1.485-2.311) | .000 |
| 18605-49730 | 3.021 (2.448-3.728) | .000 |
| ≥ 49731 | Reference |  |
| History of Comorbidity in Family |  |  |
| Present | .882 (0.723-1.075) | .212 |
| Absent | Reference |  |
| History of infection with covid in family |  |  |
| Yes | 1.595 (1.432-1.821) | .002 |
| No | Reference |  |
| Received Covid-19 vaccination |  |  |
| Yes | 1.014 (0.890-1.154) | .835 |
| No | Reference |  |
| Children Received Childhood Vaccination under NIS |  |  |
| Yes | 1.251 (1.218-1.289) | .000 |
| No | Reference |  |

Parents/ caregivers with a history of COVID-19 infection in the family demonstrated increased intent to vaccinate (OR=1.595, 95% CI [1.432-1.821], p=0.002). Children who received childhood vaccinations under the National Immunization Schedule (NIS) positively influenced parent/caregiver intent to vaccinate against COVID-19 (OR=1.251, 95% CI [1.218-1.289], p<0.001). These findings emphasize the complex interplay of behavioral and social factors in influencing parent/caregiver intentions to vaccinate against COVID-19.

### Qualitative

In total, forty-five interviews were conducted and recruitment stopped after that as data saturation was achieved. All participants were domiciled in Bihar, within the age range of 26- 64 years, two-thirds were male, with the highest level of education being graduate, majority worked in private sector, were married and belonged to Hinduism. Of all the 45 participants interviewed,28 had intention to vaccinate their children, 10 refused to get their children vaccinated against COVID-19 disease, and 7 were hesitant/unsure about COVID-19 vaccination for their children.

Figure 1 identified the facilitating and barrier factors influencing parents/caregivers’ intention to vaccinate their children against COVID-19 disease. Based on quotations from participants who intended to get their children vaccinated, the majority responded that COVID-19 vaccines are effective and would increase immunity, risk of acquiring infection is high, to protect people around and to develop herd immunity, recommended by Ministry of Health/Doctors, benefit outweighs risk and to protect children from severity/high risk. The majority of responses from almost all respondents who refused COVID-19 vaccine for their children complained about the inadequate data about the vaccine. Other reasons were related to adverse effects, beliefs, safety and inconvenience. The participants who were unsure/hesitant about COVID-19 vaccination for their children, were concerned about vaccine effectiveness, safety of vaccine, long term effect of vaccine and short duration of testing.

**Figure 1:**
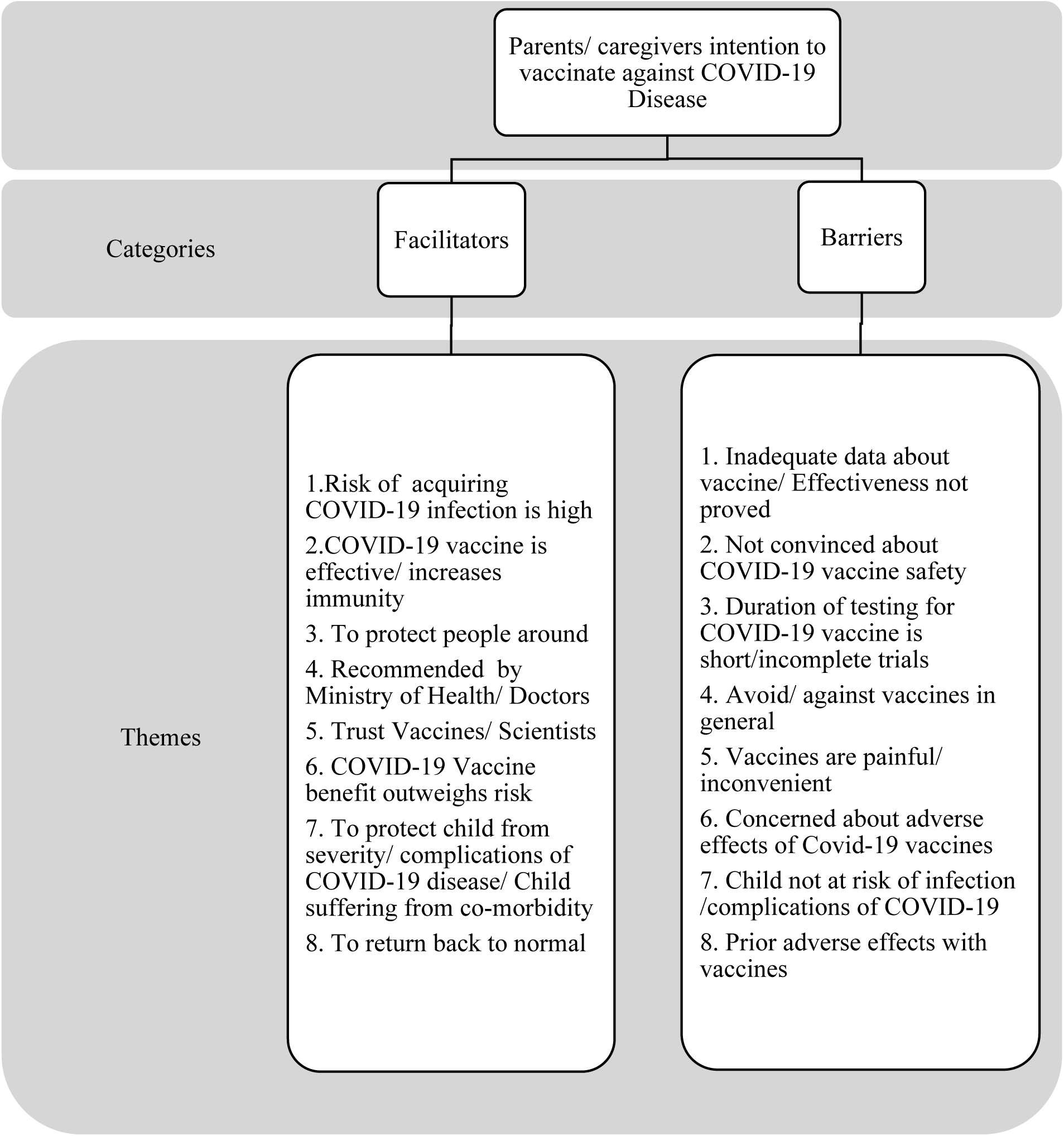
Joint display of facilitators and barriers influencing parents/caregivers’ intention to vaccinate their children against COVID-19 disease.

**Figure 2:**
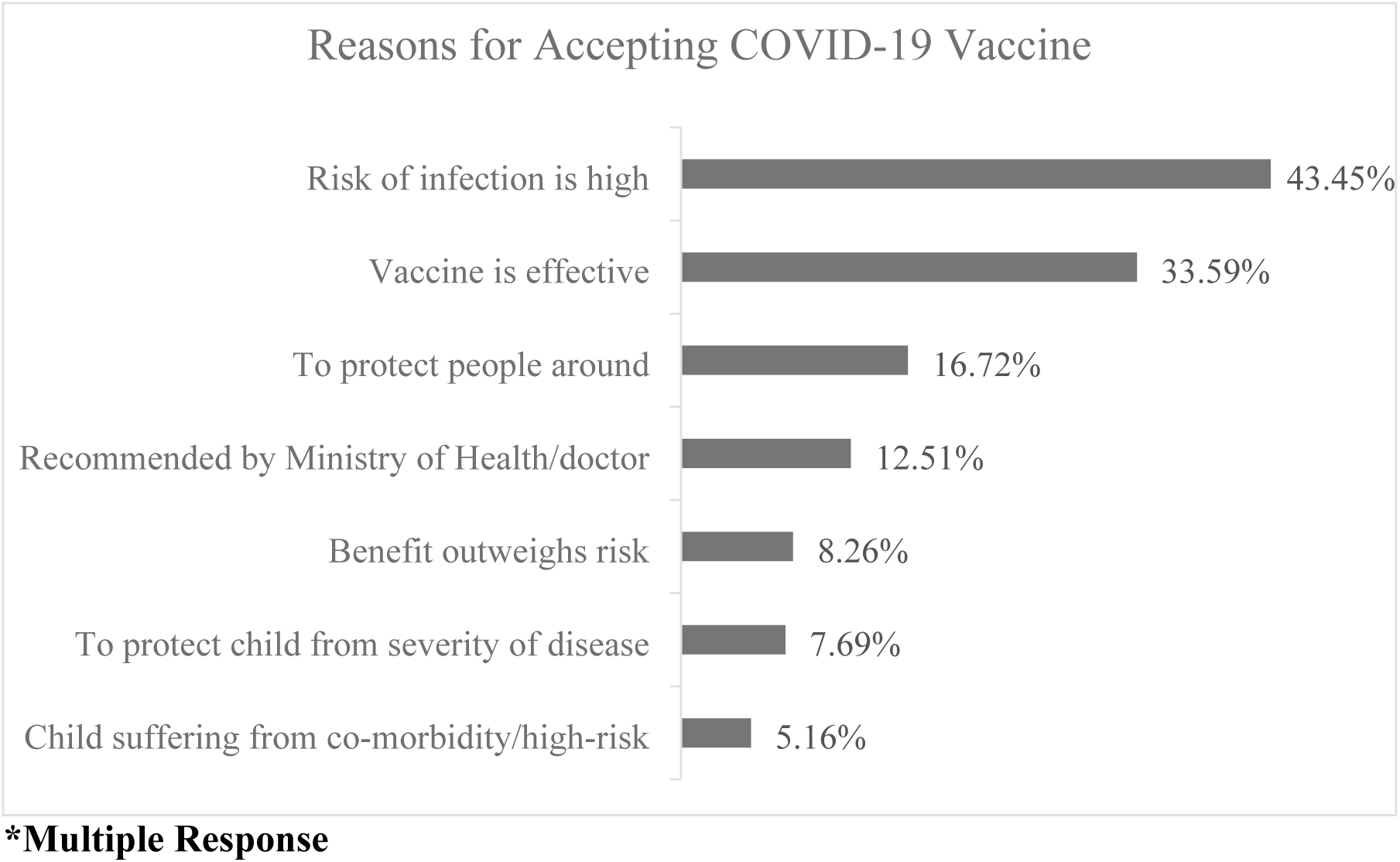
Summarizing Reasons for intending to accept COVID-19 Vaccine for children.

**Figure 3:**
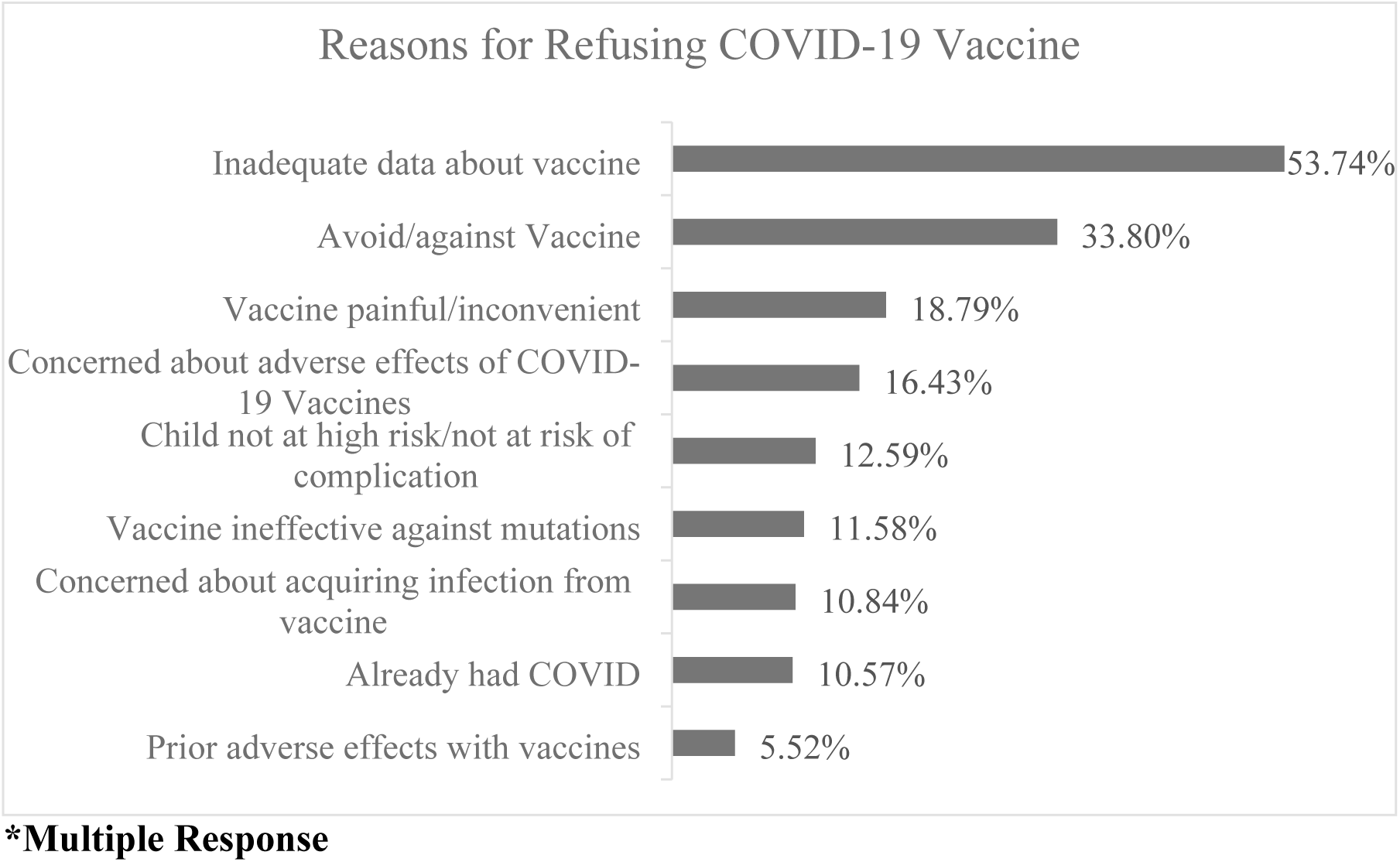
Summarizing Reasons for intending to refuse COVID-19 Vaccine for children.

**Figure 4:**
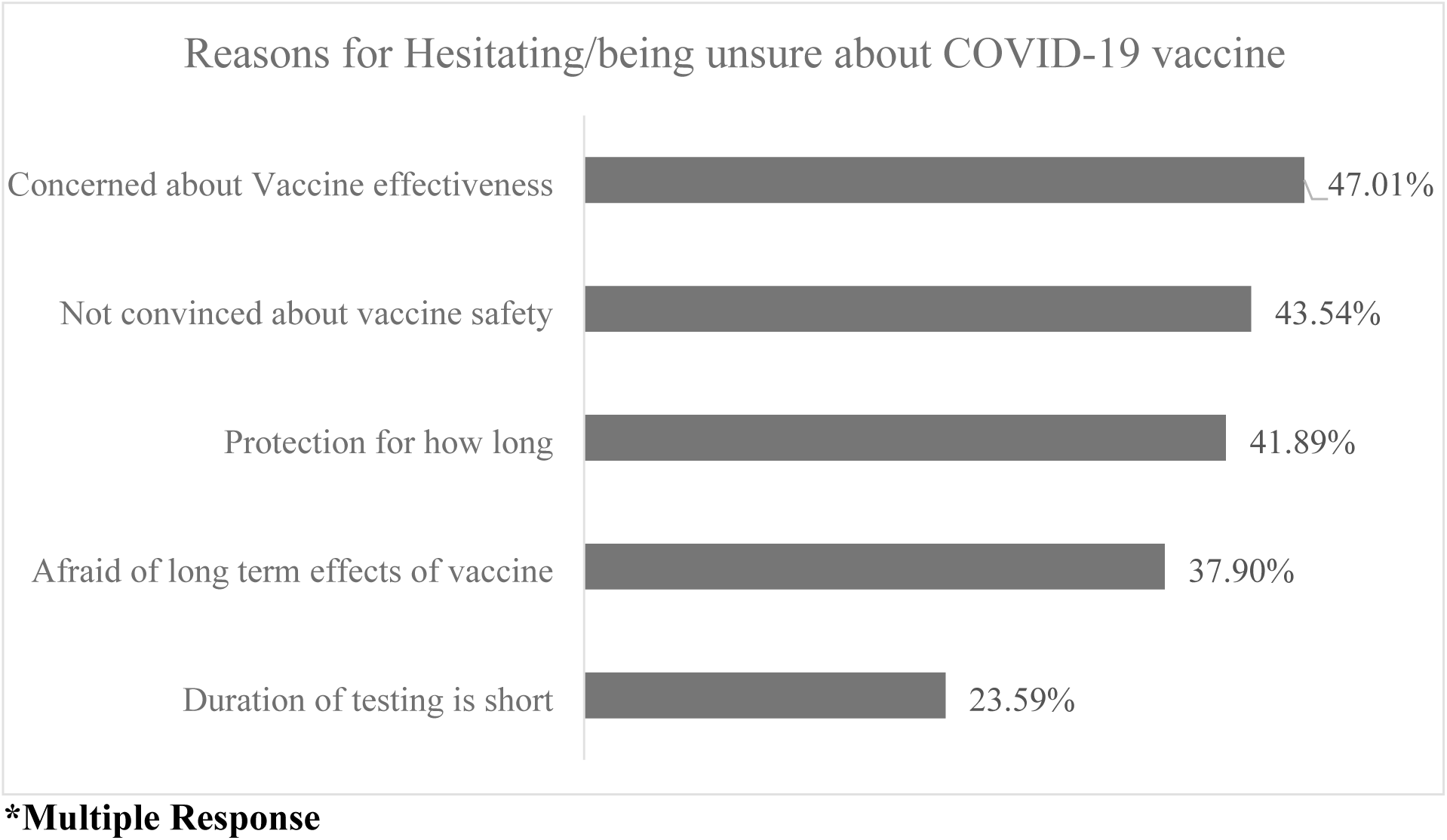
Summarizing Reasons for hesitating/being unsure about COVID-19 Vaccine for children.

### Quotes regarding intention to vaccinate against COVID-19 (Verbatim)

1. **Risk of acquiring COVID-19 infection is high-** It was perceived by many interviewees that the vaccine will protect the children from serious disease and complications of COVID-19. “If we have any underlying medical issues that could make our immune system more susceptible to infection, the risk of catching the coronavirus would increase.” (KII-07)
2. **COVID-19 vaccine is effective/ increases immunity:** “The vaccine is effective against the virus and in stopping the spread of disease, and will help to get rid of Covid-19.” (KII-30) Parents/caregivers mentioned about the importance of vaccine in achieving herd immunity, reducing the severity of COVID-19 symptoms and stopping the spread of the disease and vaccine effectiveness was an important facilitator for parents’ intention to vaccinate.
3. **To protect people around:** Parents/caregivers reason for getting vaccine themselves was to protect self and family. One of the parent’s said “I have taken the vaccine to protect our community…I am aware that the vaccine is important to protect people with co-morbidities and those with weakened immunity as they would have severe manifestations and complications of the disease.” (KII-22)
4. **Recommended by Ministry of Health/ Doctors:** Parents/caregivers will heed the advice of the medical experts and regulatory bodies tasked with weighing the risks and benefits. “It is doctors and government’s responsibility to protect us and our families, and they are encouraging us to obtain the vaccine. Since they are knowledgeable and, experts we should heed their recommendations.” (KII-12)
5. **Trust Vaccines/ Scientists:** “If there is a problem of electricity in your house, do you trust the electrician or not? Or the plumber when there’s a water problem?” …Then trust the doctors and scientists about covid!” (KII-04) “I trust the vaccine and advice given by my doctor friend and he told vaccination is important for us, so I took it… and will get it for children when it is available**”** (KII- 09)
6. **COVID-19 Vaccine benefit outweighs risk:** Parents/caregivers are making an important decision for their children based on the information available in present scenario and vaccine benefit outweighs risk. One of the parent’s said “it is a fear-based decision and risk of acquiring Covid-19 infection is high as compared to risks associated with vaccine.” (KII-17)
7. **To protect child from severity/ complications of COVID-19 disease/ Child suffering from co-morbidity:** Parents/caregivers would vaccinate their children to protect them from severe forms and complications of Covid-19 disease and also parents whose children were suffering from co-morbidity were more willing to get their children vaccinated. “My child has had all of the childhood vaccines and I would 100% vaccinate my child as soon as it is available to protect him from coronavirus.” (KII-41)
8. **To return back to normal:** The vaccination was also identified as playing an important role in returning to normalcy by many participants. In order to avoid lockdowns, resume travel, as well as for starting schools, parents said they would consent to vaccination. “It’s the only way we can hope to start our lives again without experiencing excess of cases like other countries.” (KII-24)
9. **Inadequate data about vaccine/ Effectiveness not proved:** Through our qualitative analysis we were able to get detailed concerns of parents and their reasons for refusal and being unsure about vaccine. Many parents were concerned about vaccine effectiveness, newness of vaccine, inadequate data available about vaccine and guidelines about children’s vaccination. “I don’t want to make them (children) guinea pigs…I would require more scientific and logical data before making the final decision.” (KII-02)
10. **Not convinced about COVID-19 vaccine safety**: Safety was deemed to be an important concern with the COVID-19 vaccination by majority of parents/caregivers. “I don’t trust WHO and pharmaceutical companies who put profit before lives… these vaccines are only for money making and not at all safe.” (KII-28)
11. **Duration of testing for COVID-19 vaccine is short/incomplete trials:** “I am hesitant to take a vaccine or have my child vaccinated with a vaccine whose duration of testing is too short to account for all potential long-term consequences.” (KII-05)
12. **Avoid/ against vaccines in general:** “Nothing will affect or alter my stance towards vaccination. I won’t immunize my family members. It’s my decision.” (KII-16)
13. **Vaccines are painful/ inconvenient:** Few parents gave reasons “A child should not get too many injections; it is difficult to see then in so much pain and discomfort.” (KII- 34)
14. **Concerned about adverse effects of Covid-19 vaccines:** Parents/caregivers fear of side effects was an important drivers of vaccine hesitancy. “I am worried about the side effects that will come in the years to come… maybe we should wait and protect them from potential side effects.” (KII-01)
15. **Child not at risk of infection /complications of COVID-19:** ‘Children are least likely to have it, (they) are strong against this flu, so it’s useless for them. why vaccinate (them)?” (KII-08)
16. **Prior adverse effects with vaccines:** Some parents claimed that if the child gets sick after vaccination, it also affects their attitude towards vaccination. “My neighbor’s child was serious after vaccination and was sick for many days, they had to take her to private doctor for treatment… my child also gets sick easily, so I don’t vaccinate her now.” (KII-13)

## Discussion

This was a sequential explanatory mixed-method study, with cross-sectional questionnaire-based survey to determine the prevalence of intention to vaccinate among parents/caregivers for pediatric COVID-19 vaccination, followed by semi-structured interviews to explore the facilitating and barrier factors. The data from our extensive study on parents/caregivers provide insights into the perspectives regarding COVID-19 vaccination for their children. Around 15% answered no and a little more than one-tenth (11.6%) of our participants were hesitant/unsure whether they would get their children vaccinated against COVID-19 disease. This emphasizes the significance of using open-ended questions instead of relying solely on straightforward ’yes’ or ’no’ inquiries. Our study features the significance of going past basic ’yes’ and ’no’ questions. Our quantitative analysis revealed limited reliable indicators of intention to vaccinate, but our qualitative data provided in-depth insights into parents’ intentions, yielding valuable information.

This study showed the intention to vaccinate children against COVID-19 disease was 73.4%. The acceptance rate of childhood Covid-19 vaccination has been diverse across countries, ranging from 48%to >90%^25–27^. This variation may be attributed to parents/caregivers’ perception regarding Covid vaccination, severity of the COVID-19 pandemic across countries, differences in the regulatory standards and previous adverse effects associated with childhood vaccination.

The findings of our study revealed that parent/caregiver’s education, occupation, childhood vaccination status, household suffering with chronic diseases and previous COVID-19 infection in family were significantly associated with parent/caregiver’s intention to vaccinate against COVID-19 disease for their children. Similar findings were also observed in studies carried out in other regions.^28,29^

In our study, the major reasons for accepting COVID-19 vaccination were due to high risk of infection to child, belief in vaccine being effective, to protect people around/develop herd immunity, benefit outweighs risk.

The major reasons for refusing COVID-19 vaccination were inadequate data about vaccine, against vaccine in general, vaccine inconvenient, concerned about side effects or acquiring disease from the vaccine, and previous adverse effect with vaccine.

The major reasons for hesitating about COVID-19 disease were concerns about vaccine effectiveness, not convinced about vaccine safety, concerns about how long the protection will last, and afraid of long-term effects of vaccine.

The strength of our study was that it adopted a sequential explanatory mixed method design and explored reasons for accepting, refusing, as well as hesitancy regarding Covid-19 vaccination for children. Thus, we were able to acquire a plethora of information by allowing participants to express their ideas and concerns in their own words, which would not have been possible with only closed-ended questions. A large representative sample of parents from the region participated in the study and were interviewed regarding their perceptions and intentions to vaccinate their children. The robustness of our procedures and the saturation of the topics after 45 interviews both attest to the internal validity of the study The study had intrinsic limitations. Despite using both online and offline questionnaires which was useful for data collection, our sampling strategy might not be representative of all parents in the region thus limiting the generalizability of our finding. We used mobile phones for in-depth interviews to allow participation during the Covid-19 pandemic as face-to-face interviews were not possible. This might have hampered the rapport between the interviewer and participants. Also, social desirability bias might have made it harder for people to express their true intentions regarding vaccination at the time of COVID-19 pandemic. In order to address these issues, we used a friendly, empathetic tone and interviewing style and gave sufficient time to let participants speak at their own pace, and appreciated their open responses. Further prospective multi-centered studies should be conducted in a larger population to increase the generalizability.

## Conclusion

The study adopted sequential explanatory mixed methods and thus provides a more comprehensive data, as it explored reasons for refusal against COVID-19 vaccination to facilitate recommendations to increase the rate of vaccination among children. Findings from the quantitative and qualitative analysis of our study suggest that parents/caregivers belonging to higher age group, graduate and above education group and private sector employees were less likely to have intentions towards vaccinating their child against COVID-19. Also, a history of COVID-19 infection in family and those children who received childhood vaccination their parents/ caregivers demonstrated increased intent to vaccinate. Majority of parents/caregivers had concerns around safety and efficacy of the newly developed vaccines.

To enhance the acceptance of COVID-19 vaccines among children, addressing parental concerns related to safety, efficacy, and testing emerges as a crucial factor. Implementing targeted public health strategies that involve transparent communication regarding the safety and effectiveness of vaccines can potentially enhance acceptance rates for pediatric COVID- 19 vaccination.

## Data Availability

All relevant data are within the manuscript and its Supporting Information files.

## Acknowledgments

This research was funded by the Indian Council of Medical Research (ICMR), Department of health Research, Ministry of Health and Family Welfare, Government of India. The authors acknowledge Smt. Shefali Kuntal, Shri Sanjeet Kumar, Shri Suman Kumar, Shri Manish Kumar and other staff for technical support in data collection and data analysis. Also, the authors acknowledge all senior residents, junior residents and participants who participated in this study.

